# Context-Dependent FHIR Serialisation Strategies for Clinical LLM Deployment: A Multi-Layer Benchmark on UK Core Data

**DOI:** 10.64898/2026.08.05.26359794

**Authors:** Jacqueline Chong

## Abstract

The choice of FHIR-to-text serialisation format significantly impacts clinical LLM quality (Kruskal–Wallis H=163.86, p< 10^−33^, Δ=0.24 on a 5-point scale), yet remains unstudied as a clinical deployment variable. We present FHIRBench-UK, evaluating five large language models across six serialisation formats and three clinical tasks on 100 UK Core FHIR patient bundles (18,000 scored prompts across clean and perturbed cohorts). The optimal format is context-dependent: raw_json dominates for clinical QA, hybrid_adaptive for clinical reasoning, and structured_markdown for summarisation. In 58% of model–task–complexity scenarios, raw_json is suboptimal. Model capability moderates format sensitivity: Claude Sonnet 4.5 shows 0.10-point sensitivity versus Llama 3.3’s 0.39, making adaptive serialisation most valuable for budget-constrained deployments using mid-tier models. All findings replicate under clinically realistic data perturbation. The study additionally confirms a complete ranking inversion between token-level F1 and clinical quality (*ρ*=−0.90), replicating US findings across UK Core profiles, and converges with independent work on open-weight models [1] to establish serialisation strategy as a replicable determinant of clinical LLM performance. We recommend task-aware serialisation routing as a zero-cost quality intervention for NHS FHIR-based LLM deployments.

## 1 Introduction

NHS England’s 10-Year Health Plan [2] commits to artificial intelligence–enabled clinical workflows across primary, secondary, and community care settings. The technical infrastructure underpinning this ambition increasingly centres on FHIR (Fast Healthcare Interoperability Resources) [3], now mandated for all new NHS APIs—with over 187 FHIR-based APIs in the NHS API catalogue as of 2026 [4, 5]. As NHS trusts, integrated care systems, and GP federations adopt FHIR-based interoperability standards, the volume of structured clinical data available for AI processing grows rapidly. Large language models are being evaluated and deployed for clinical tasks against this data: extracting information from patient records, reasoning about drug interactions, generating discharge summaries, supporting clinical coding, and assisting clinical decision-making [6, 7]. Yet a critical preprocessing step in every such deployment—how structured FHIR data is converted to text for LLM consumption—remains entirely unstudied as a clinical quality variable. This study addresses that gap.

FHIR resources are stored as hierarchical JSON structures conforming to national profiles (UK Core in England and Wales) [8, 9]. Before an LLM can process a patient bundle, the structured JSON must be serialised into a text representation that fits within the model’s context window. Multiple serialisation strategies exist: passing the raw JSON verbatim, flattening the hierarchy into dot-notation key-value pairs, converting to clinical narrative prose, rendering as structured markdown with headings, or applying domain-specific clinical templates that reorganise information by clinical category. Each format makes different tradeoffs. Raw JSON preserves all structural information but consumes 5–10× more tokens than alternatives. Narrative formats achieve substantial token reduction but may omit system URIs and coded identifiers. Template formats pre-organise clinical concepts but impose assumptions about information priority. The choice of serialisation format thus determines three deployment-critical variables simultaneously: what clinical information the LLM receives, how that information is semantically organised, and how much the inference costs. For a typical complex patient bundle (80+ FHIR resources), raw JSON may consume 10,000 input tokens whilst a clinical template representation of the same data requires fewer than 1,000—a 10× cost differential at scale. Every deployment that connects a FHIR data source to an LLM implicitly makes a serialisation choice—the model receives text, not structured objects. In practice, most implementations default to passing raw JSON without considering alternatives; serialisation is treated as a transparent implementation detail rather than a design decision with quality consequences. This study demonstrates that it is the latter.

Our previous work (FHIRBench [10]) provided initial evidence that serialisation format affects LLM performance on US Core FHIR data. That study evaluated five models across multiple serialisation formats on token-level F1 (Layer 1) and LLM-as-judge clinical quality (Layer 2), identifying a striking ranking reversal between layers—models that excelled on automated metrics performed worst on clinical quality, and vice versa. However, its analysis of serialiser effects was limited in three respects. First, it assessed serialiser impact primarily through automated metrics rather than clinical quality dimensions. Second, it used a single data condition without perturbation robustness testing, leaving open whether observed effects would hold under real-world data noise. Third, it evaluated US Core FHIR profiles; generalisability to UK Core—with its distinct terminology systems (SNOMED CT UK Edition [11], dm+d for medications [12]), mandatory extensions (NHS Number verification status, ethnic category), and clinical documentation conventions—remained unestablished. The present study addresses all three limitations.

Concurrent with our Paper 1, Pator (2026) [1] presented the first systematic comparison of FHIR serialisation formats for a single clinical task (medication reconciliation), demonstrating a 19 F1-point advantage for clinical narrative over raw JSON in sub-8B open-weight models—an advantage that reversed for the largest model tested (Llama 3.3 70B, where raw JSON achieved F1=0.996). That study evaluated four formats across five open-weight models on 200 synthetic patients using F1 scoring alone. The present study extends Pator’s findings in five directions: multi-task evaluation (QA, reasoning, summarisation), clinical quality assessment beyond F1, UK Core FHIR profiles, perturbation robustness testing, and frontier proprietary models (Claude, GPT-5.4). Critically, the two studies—conducted independently using different models, tasks, and data—converge on the same core conclusion: serialisation strategy significantly impacts clinical LLM performance, and the optimal format is context-dependent. This convergent evidence from independent conditions strengthens the claim beyond what either study alone could establish. A growing ecosystem of FHIR-LLM benchmarks—including FHIR-AgentBench [13], FHIR-AgentEval [14], MedA-gentBench [15], and LLM FHIR Eval [16]—further confirms that the FHIR-to-LLM interface is emerging as a critical research frontier, though none of these address the serialisation format question directly.

The present study makes three contributions that advance the field beyond Paper 1:

1. **First comprehensive evaluation of serialisation strategy impact on clinical quality for UK Core FHIR data.** We demonstrate that format choice significantly affects clinical quality—an effect previously undetected due to evaluation methodology limitations that restricted analysis to the model least sensitive to format variation.
2. **Discovery of context-dependent serialiser optimality.** The optimal serialisation format varies by clinical task—supporting task-adaptive serialisation rather than static format selection. In the majority of evaluation scenarios, raw_json is suboptimal; alternative formats outperform it substantially in specific task–complexity combinations. This finding motivates a zero-cost quality intervention requiring only preprocessing routing, no model retraining or prompt engineering.
3. **Perturbation robustness across two independent cohorts.** All findings—serialiser rankings, task-specific optimality, model–serialiser interactions, and the ranking reversal between evaluation layers—are robust under clinically realistic data perturbation, establishing that the reported effects are intrinsic properties of the format representations rather than artefacts of idealised synthetic data.

These contributions are evaluated through FHIRBench-UK: 100 UK Core FHIR patient bundles stratified by clinical domain and complexity, processed through six serialisation formats across three clinical tasks by five LLMs (Table 1: Claude Sonnet 4.5, GPT-5.4, DeepSeek V3.2 [17], Qwen3 32B, Llama 3.3 70B), yielding 18,000 scored prompts per evaluation layer under two data conditions. The benchmark, evaluation pipeline, and complete dataset are released as open resources to support reproducibility and community extension.

**Table 1:**
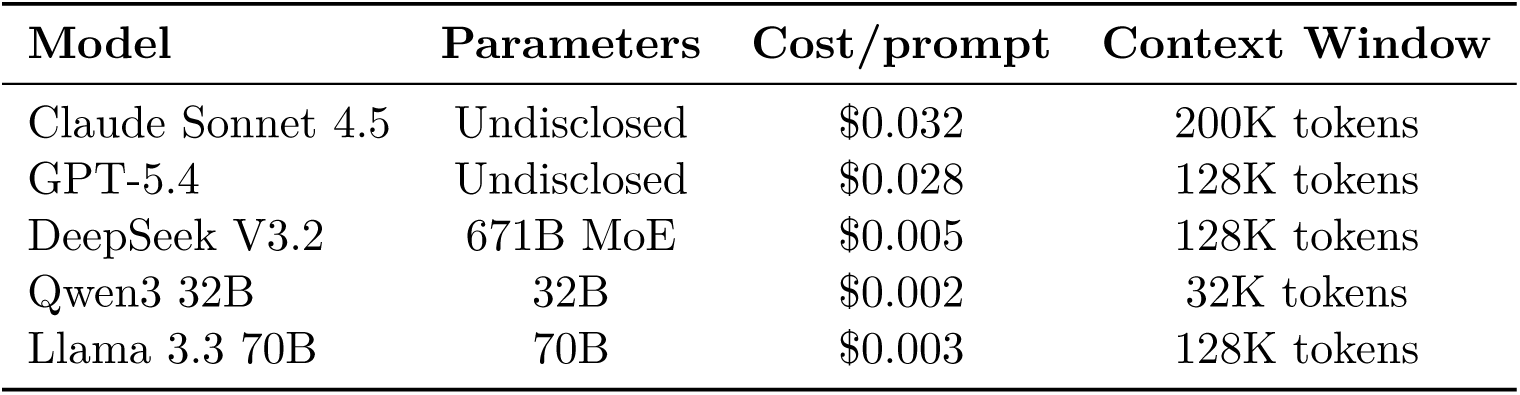
Models under evaluation. All accessed via AWS Bedrock with temperature=0.0.

The paper is structured as follows. §2 describes the evaluation cohort, serialisation formats, clinical tasks, experimental design, and perturbation protocol. §3 presents results with serialisation strategy as the primary analytical lens. §4 discusses mechanisms, NHS deployment implications, and the adaptive serialisation case. §5 concludes with recommendations for clinical AI practitioners.

## 2 Methods

### 2.1 Evaluation Cohort

The evaluation cohort comprises 100 UK Core FHIR R4 patient bundles, stratified-sampled from a pool of 992 generated patients. Each bundle represents a complete primary care patient record conforming to NHS England’s UK Core Implementation Guide (v1.0.0) [8], containing Patient, Condition, MedicationRequest, Observation, Organization, and Practitioner resources with appropriate inter-resource references.

#### 2.1.1 Generation Approach

Patient bundles were generated using DeepSeek V3.2 [17] (open-weight, 671B MoE architecture) via AWS Bedrock. We selected LLM generation over the established Synthea synthetic patient generator [18] for seven specific reasons related to UK Core compliance:

1. **dm+d medication codes**: Synthea uses US RxNorm codes. UK Core mandates Dictionary of Medicines and Devices (dm+d) [12,19,20] codes for all MedicationRequest resources. No maintained mapping exists between RxNorm and dm+d.
2. **NHS Number format**: UK Core requires 10-digit NHS Numbers validated by the Modulus 11 check digit algorithm. Synthea generates US SSN-format identifiers.
3. **UK Core extensions**: Mandatory extensions including *UKCoreEthnicCategory*, *UKCoreNHSNumber-VerificationStatus*, and *UKCoreResidentialStatus* have no Synthea equivalent.
4. **GP practice registration**: UK primary care records include explicit `generalPractitioner` references with ODS codes. Synthea models US provider networks.
5. **SNOMED CT UK Edition**: UK Core requires codes from the UK clinical extension of SNOMED CT [11], which includes ∼50,000 additional concepts not in the International Edition.
6. **Metric units**: UK clinical observations use metric units exclusively (mmol/L for glucose, mmol/-mol for HbA1c, mmHg for blood pressure). Synthea defaults to US conventional units for several observations.
7. **Bundle structure conventions**: UK Core bundles follow NHS Digital’s structure guidance for document bundles [5], with specific ordering and reference patterns.

Generation was configured with `max_tokens=32768`, `temperature=0.0` (for reproducibility), `read_timeout=600s`, and 5 concurrent workers. Each patient was generated with a structured prompt specifying clinical domain, complexity level, demographic characteristics, and required UK Core conformance criteria.

#### 2.1.2 Stratification

The 100-patient evaluation cohort was stratified-sampled from the 992-patient pool along two dimensions:

**Complexity** (determined by resource count and clinical scenario):

- Simple (25 patients): 10–20 resources. Single chronic condition, 1–2 medications, routine observations.
- Moderate (40 patients): 20–40 resources. 2–3 conditions, polypharmacy (3–5 medications), multiple observation types.
- Complex (25 patients): 40–70 resources. 4+ conditions, significant polypharmacy, multi-year observation history, specialist referrals.
- Highly complex (10 patients): 70+ resources. Multi-morbidity with interactions, complex medication regimens, extensive longitudinal data.

##### Clinical domain

Diabetes (24%), Cardiovascular (31%), Preventive care (28%), Medication interactions (17%). Sampling used a fixed random seed (42) with domain balance constraints.

#### 2.1.3 Validation

All 100 bundles achieved 100% pass rate on seven mandatory UK Core conformance criteria: Bundle structure, NHS Number (Modulus 11 validated), dm+d codes [21, 22], SNOMED CT UK codes, UK Core extensions, GP practice reference, and metric units. Validation was performed programmatically using `validate_uk_core.py`.

### 2.2 Serialisation Formats

Six serialisation formats convert each FHIR Bundle JSON into text for LLM consumption. This represents the most comprehensive format comparison to date; Pator (2026) [1] evaluated four formats on a single task, whilst our design crosses six formats with three tasks, enabling discovery of task-specific optimality. All serialisers are deterministic and implemented as Python classes.

#### 2.2.1 raw_json

The complete FHIR Bundle JSON is passed as a formatted string with 2-space indentation. All structural information is preserved. Mean 9,901 input tokens per prompt.

#### 2.2.2 flattened_kv

The JSON hierarchy is flattened into dot-notation key-value pairs. Each leaf value receives a fully-qualified path key. Mean 6,081 input tokens (39% reduction vs raw_json). Preserves all data but fragments clinical concepts across disconnected lines.

#### 2.2.3 narrative

Converts the FHIR Bundle into NHS clinical letter format—GP referral style prose. Mean 1,179 input tokens (88% reduction). Presents information in a format familiar to clinicians.

#### 2.2.4 clinical_template

Reorganises bundle content into an NHS SOAP-style clinical template with standardised sections. Mean 942 input tokens (91% reduction). Uses clinical abbreviations and tabular layout.

#### 2.2.5 structured_markdown

Renders the bundle as hierarchical markdown with headings, subheadings, and nested lists. Mean 1,226 input tokens (88% reduction). Preserves hierarchical document structure via heading levels.

#### 2.2.6 hybrid_adaptive

A task-aware meta-serialiser that selects format based on the clinical task: clinical_qa → clinical_template; clinical_reasoning → structured_markdown; clinical_summarization → narrative. Mean 1,116 input tokens (89% reduction).

### 2.3 Clinical Tasks

Three clinical tasks span the range of NHS primary care AI applications:

#### 2.3.1 Clinical QA (Factual Extraction)

The model must extract five specific clinical facts: NHS Number, active conditions with SNOMED CT UK codes, current medications with dm+d codes, most recent HbA1c result, and registered GP practice.

#### 2.3.2 Clinical Reasoning

The model must identify potential drug interactions or contraindications, assess concerning trends in observations, recommend clinical actions, and identify care plan gaps.

#### 2.3.3 Clinical Summarisation

The model must produce a comprehensive GP referral letter including demographics, conditions with codes, medications with doses, investigation results with trends, relevant history, care plan, and referral rationale.

### 2.4 Scoring Methodology

#### 2.4.1 Layer 1: Token-Level F1

Ground truth is extracted programmatically from each FHIR Bundle. Model responses are tokenised (lowercased, split on non-alphanumeric characters, stopwords removed), and token-level precision, recall, and F1 are computed. Patient-level aggregation (N=100) provides the unit of analysis.

#### 2.4.2 Layer 2: LLM-as-Judge Clinical Quality

A cross-judging protocol mitigates self-assessment bias [23]: Claude Sonnet 4.5 judges responses from GPT-5.4, DeepSeek V3.2, Qwen3 32B, and Llama 3.3 70B (7,200 judgments); Qwen3 32B judges Claude responses (1,800 judgments). Each judgment scores on four dimensions (0–5): Accuracy, Completeness, Safety, and Relevance. Temperature is 0.0 for reproducibility.

#### 2.4.3 Statistical Tests

Inter-model and inter-serialiser differences are assessed using the Kruskal–Wallis H-test. Post-hoc pairwise comparisons use Dunn’s test with Bonferroni correction. Effect sizes use Cohen’s d. All confidence intervals are 95%.

### 2.5 Models Under Evaluation

Five large language models were evaluated, representing the major architecture families available through AWS Bedrock as of July 2026:

All models were accessed via AWS Bedrock with shared configuration: `temperature:` 0.0, `max_tokens:` 4096, `read_timeout:` 600 seconds, concurrency: 5 workers per model. GPT-5.4 was accessed via Bedrock’s Mantle integration (OpenAI Responses API). No system prompts were used.

### 2.6 Prompt Generation

The full evaluation matrix is:

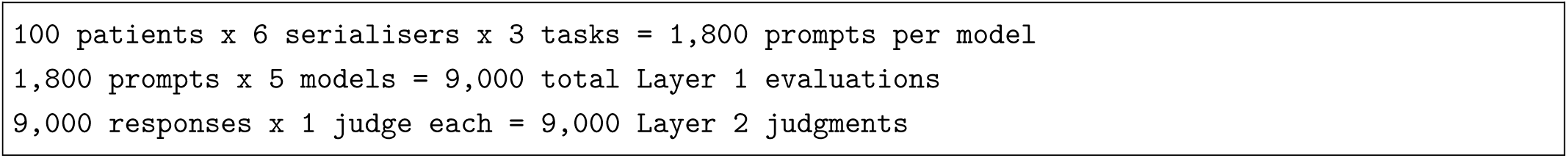

Each prompt is constructed by loading the patient FHIR Bundle JSON, applying the specified serialiser, inserting the serialised text into the task-specific prompt template, and recording metadata. The prompt set is fixed across all models. All five models achieved ≥99.8% success rate across both cohorts.

### 2.7 Perturbation Protocol

The perturbation experiment applies clinically realistic data noise to the evaluation cohort, simulating quality issues characteristic of production NHS records. Eight perturbation types were implemented:

1. **Duplicate MedicationRequests** (34/100 patients): GP system renewal duplicates.
2. **Legacy Read codes** (21/100): Read v2 codings alongside SNOMED CT.
3. **Missing observation units** (47/100): Removed valueQuantity.unit fields.
4. **Free-text abbreviated clinical notes** (33/100): NHS clinical abbreviations.
5. **Mixed date formats** (25/100): DD/MM/YYYY, D-Mon-YY variations.
6. **Incomplete medication histories** (39/100): Medications marked as entered-in-error.
7. **Contradictory entries** (39/100): Active conditions with abatement dates.
8. **Variable terminology** (38/100): Clinical abbreviations (“T2DM”, “AF”).

Perturbation intensity scales with complexity (Simple: 1–2; Moderate: 2–3; Complex: 3–4; Highly complex: 4–5). The perturbed cohort undergoes the identical evaluation pipeline with scoring against perturbed ground truth.

### 2.8 Cost-Efficiency Analysis

Per-prompt cost is calculated as: Cost = (input_tokens*/*10^6^) × input_rate + (output_tokens*/*10^6^) × output_rate.

**Table 2:** AWS Bedrock pricing (us-east-2, July 2026). GBP figures use $1 = £0.79.

| Model | Input Rate (USD/M tokens) | Output Rate (USD/M tokens) |
| --- | --- | --- |
| Claude Sonnet 4.5 | \$3.00 | \$15.00 |
| GPT-5.4 (Mantle) | \$2.50 | \$10.00 |
| DeepSeek V3.2 | \$0.62 | \$1.85 |
| Llama 3.3 70B | \$0.72 | \$0.72 |
| Qwen3 32B | \$0.20 | \$0.78 |

### 2.9 Ethical Considerations

The study did not require ethics committee approval. The benchmark design aligns with NICE’s evidence standards framework for digital health technologies [24] and MHRA’s regulatory expectations for AI-as-medical-device validation [25].

#### Use of AI Tools

Large language models (specifically Amazon Quick, an AI assistant built on Claude) were used to assist with code generation for benchmark scripts, statistical analysis, data processing pipelines, and manuscript drafting. All AI-generated code was reviewed and validated by the author. All scientific claims, experimental design decisions, statistical interpretations, and conclusions are solely the responsibility of the author.

## 3 Results

The evaluation comprised 18,000 prompts across five models, six serialisation formats, three clinical tasks, and four complexity levels, assessed under two conditions (clean and perturbed cohorts) using both token-level F1 (Layer 1) and LLM-as-judge clinical quality scoring (Layer 2).

### 3.1 Serialisation Format Significantly Impacts Clinical Quality

The choice of FHIR-to-text serialisation format produces a clinically meaningful difference in LLM clinical quality. Across all models and tasks, Layer 2 composite scores varied by 0.24 points (Table 3) on a 5-point scale as a function of serialiser alone (Kruskal–Wallis H=163.86, df=5, p< 10^−33^).

**Table 3:**
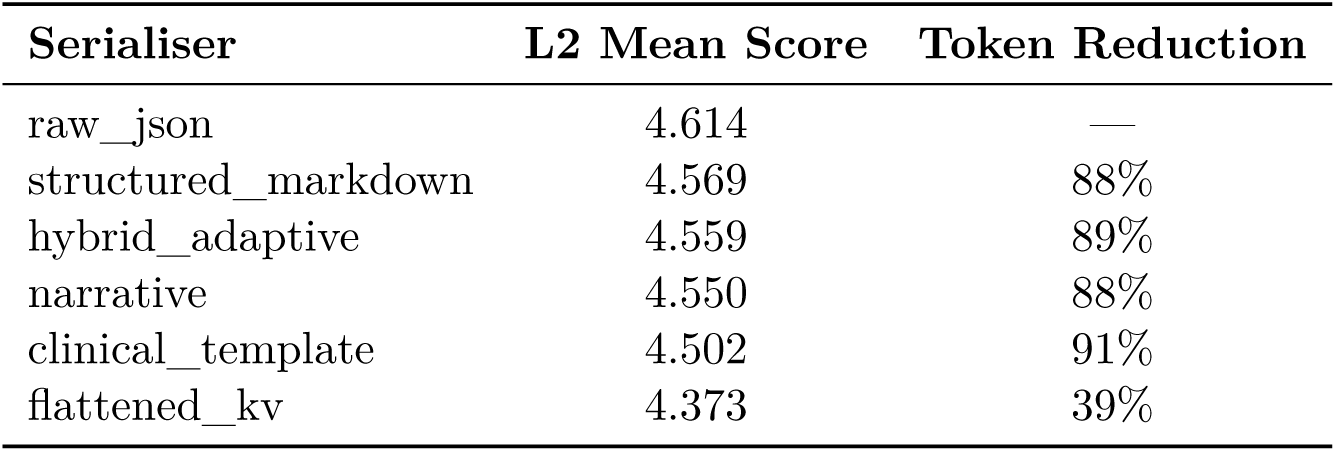
Layer 2 clinical quality by serialisation format (clean cohort, all models pooled).

On Layer 1 (token-level F1), the serialiser range was 0.042 (raw_json: 0.451, flattened_kv: 0.409). On Layer 2 under data perturbation, the effect remained significant (H=154.12, df=5, p< 10^−31^; range 0.20). The consistency across layers, cohorts, and models establishes serialisation format as a reliable, controllable variable in clinical LLM deployment.

### 3.2 The Optimal Serialiser Is Context-Dependent

The aggregate ranking conceals a critical finding: the optimal serialisation format changes depending on clinical task and data complexity (Table 4). raw_json is not universally optimal; in 58% of model–task–complexity scenarios (35 of 60), an alternative format outperforms it.

**Table 4:** Layer 2 mean score by serialiser and clinical task (all models pooled).

| Serialiser | Clinical QA | Clinical Reasoning | Clinical Summarisation |
| --- | --- | --- | --- |
| raw_json | **4.867** | 4.515 | 4.520 |
| structured_markdown | 4.733 | 4.530 | **4.518** |
| hybrid_adaptive | 4.744 | **4.576** | 4.434 |
| narrative | 4.722 | 4.543 | 4.465 |
| clinical_template | 4.668 | 4.559 | 4.356 |
| flattened_kv | 4.432 | 4.364 | 4.329 |

Three task-specific patterns emerge:

- **Clinical QA** (factual extraction): raw_json dominates. Complete preservation of identifiers, codes, and system URIs provides verbatim access to exact tokens needed.
- **Clinical reasoning** (drug interactions, risk assessment): hybrid_adaptive and clinical_template outperform raw_json. Reasoning tasks benefit from semantic organisation that groups related clinical concepts.
- **Clinical summarisation**: structured_markdown performs comparably to raw_json whilst achieving 88% token reduction.

In 17 of 60 scenarios, alternative formats beat raw_json by more than 0.1 points (Table 5); in 10, by more than 0.2. The largest reliable reversal (+0.58) demonstrates that structured_markdown can shift output from marginally acceptable (3.14) to clinically adequate (3.72) solely by changing serialisation format.

**Table 5:** Largest serialiser advantages over raw_json (model × task × complexity scenarios).

| Model | Task | Complexity | Best Serialiser | $\Delta$ vs raw_json |
| --- | --- | --- | --- | --- |
| Llama 3.3 | Summarisation | Highly complex | structured_markdown | +0.58 |
| Qwen3 | Summarisation | Highly complex | structured_markdown | +0.54 |
| Llama 3.3 | Summarisation | Simple | structured_markdown | +0.43 |
| Llama 3.3 | Summarisation | Moderate | structured_markdown | +0.42 |
| Qwen3 | Reasoning | Moderate | hybrid_adaptive | +0.39 |

### 3.3 Model Capability Moderates Serialiser Sensitivity

The serialiser effect is not uniform across models. Weaker models exhibit greater sensitivity to input format, whilst stronger models partially compensate for suboptimal serialisation.

The correlation between model overall quality and serialiser robustness is monotonic (Table 6, Figure 2): Claude (highest quality, lowest sensitivity) through to Llama (lowest quality, highest sensitivity). Figure 2 visualises this interaction.

**Table 6:** Serialiser sensitivity by model (Layer 2, clean cohort).

| Model | L2 Range | Best Format | Worst Format | Mean L2 |
| --- | --- | --- | --- | --- |
| Claude Sonnet 4.5 | 0.10 | raw_json (4.96) | clinical_template (4.86) | 4.90 |
| GPT-5.4 | 0.19 | raw_json (4.86) | flattened_kv (4.68) | 4.80 |
| DeepSeek V3.2 | 0.23 | raw_json (4.80) | flattened_kv (4.57) | 4.67 |
| Qwen3 32B | 0.29 | hybrid_adaptive (4.29) | flattened_kv (4.00) | 4.19 |
| Llama 3.3 70B | 0.39 | raw_json (4.10) | flattened_kv (3.71) | 3.96 |

### 3.4 Model Performance and the Ranking Reversal

On Layer 1 (token-level F1), Llama 3.3 ranked first (0.454) followed by Qwen3 (0.448), Claude (0.428), GPT-5.4 (0.417), and DeepSeek (0.416). On Layer 2 (clinical quality), the ranking inverted completely: Claude first (4.90), GPT-5.4 (4.80), DeepSeek (4.67), Qwen3 (4.19), Llama last (3.96). The Spearman correlation between layers was *ρ* = −0.90 (Figure 1).

**Figure 1:**
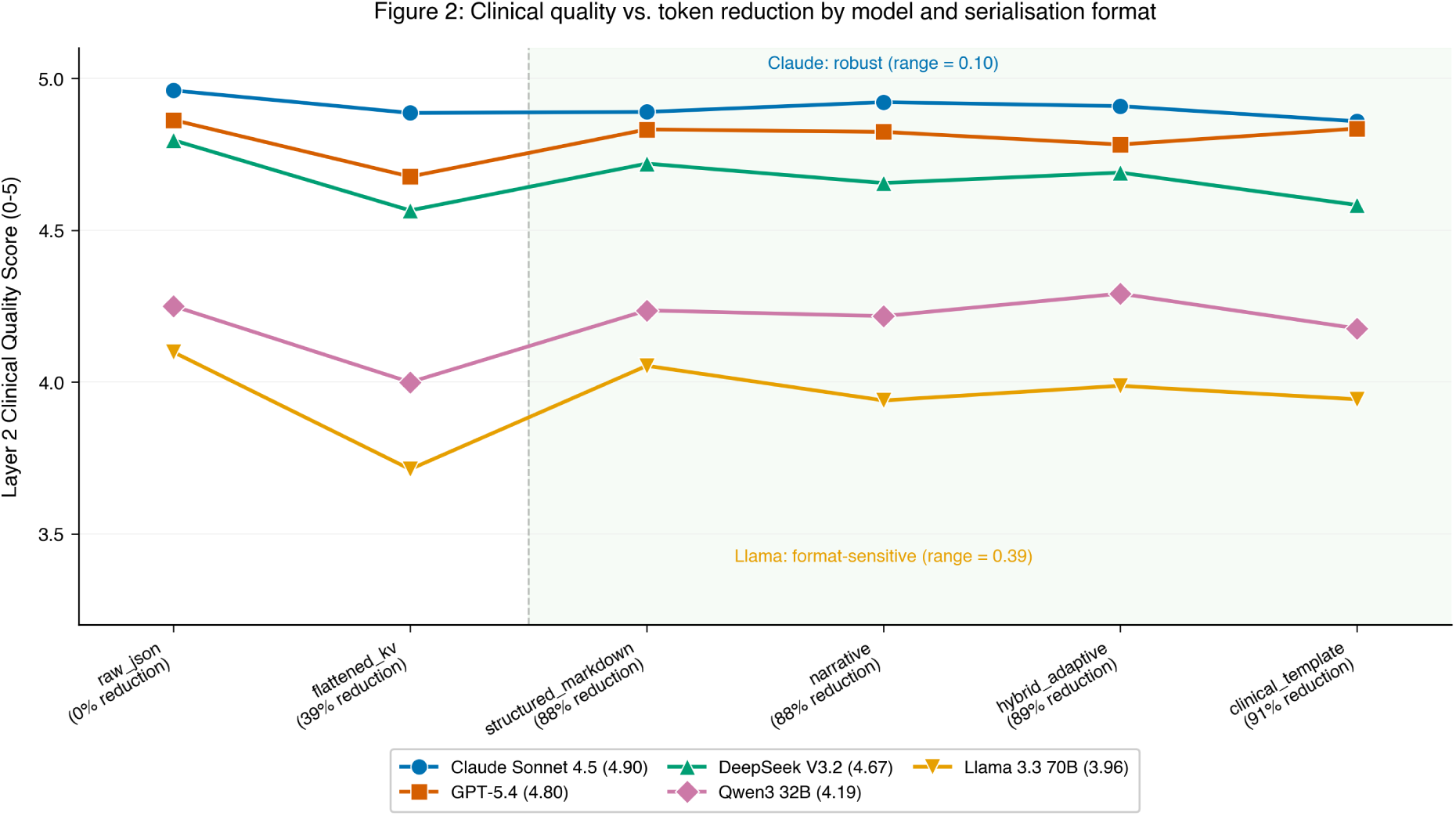
Model quality versus serialiser sensitivity. Claude’s line is nearly flat across serialisers, whilst Llama’s drops steeply—demonstrating that weaker models derive the greatest benefit from optimal format selection.

**Figure 2:**
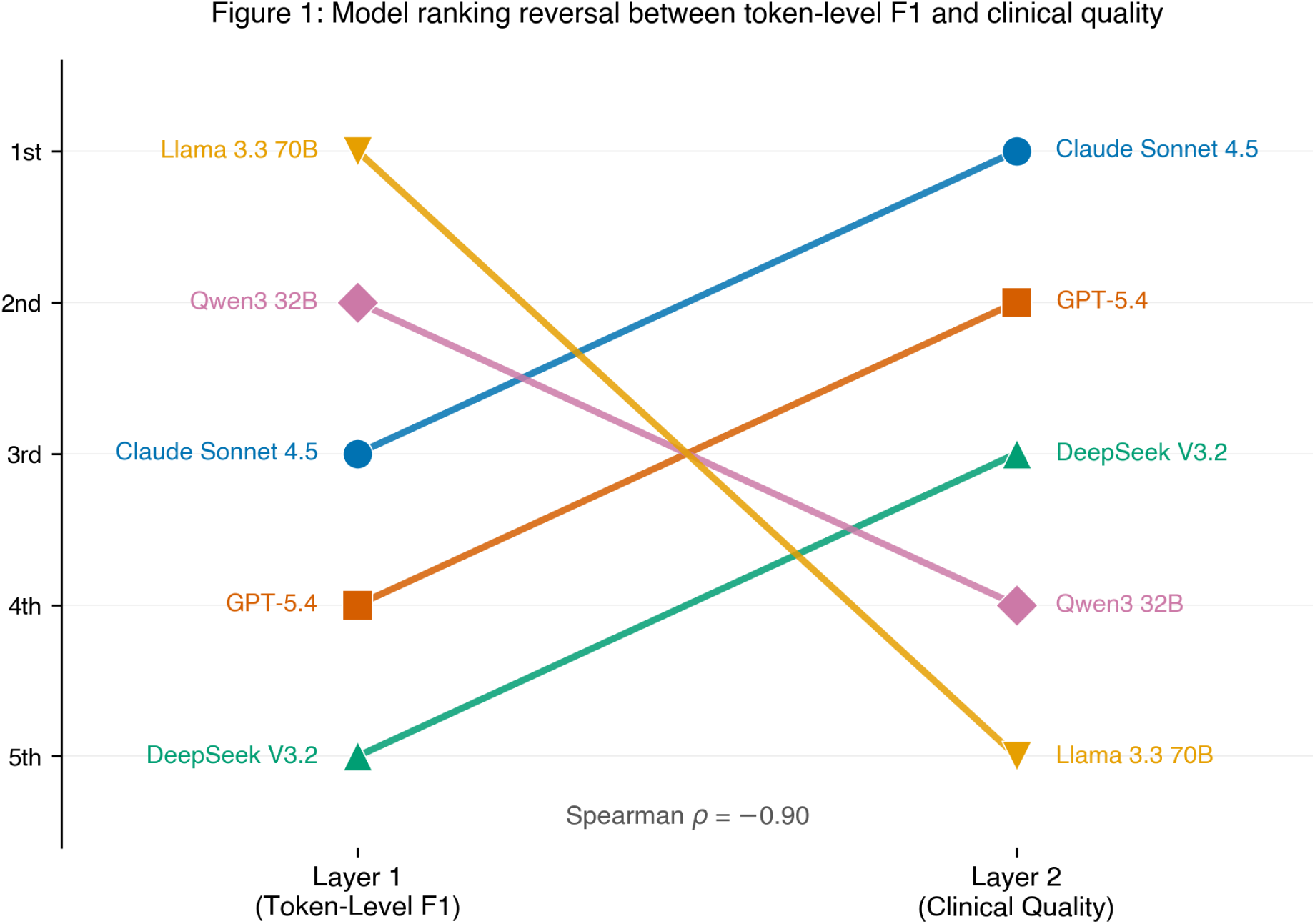
Complete ranking reversal between Layer 1 (token-level F1) and Layer 2 (clinical quality). This replicates the finding from Paper 1 [10] on US Core FHIR data.

This ranking reversal replicates the central finding of Paper 1 [10] on US Core FHIR data. Both Layer 1 and Layer 2 rankings were preserved identically on the perturbed cohort, confirming robustness to data quality variation.

### 3.5 Perturbation Robustness

Key stability findings under clinically realistic noise:

- **Model rankings**: Identical across clean and perturbed cohorts for both layers.
- **Layer 1 degradation**: Uniform −2.2% across all models (mean Δ = −0.010 F1).
- **Serialiser rankings**: Best (raw_json) and worst (flattened_kv) unchanged for all five models.
- **Serialiser effect size**: Stable between cohorts (paired t-test: t=−1.40, p=0.88).
- **Task-specific patterns**: Replicated under perturbation.

### 3.6 Cost-Efficiency Implications

Serialisation format provides a second cost lever beyond model selection (Table 2, Figure 3). Narrative and clinical_template formats achieve 88–91% input token reduction with Layer 2 quality loss of only 0.06–0.11 points versus raw_json. Combining clinical_template serialisation with DeepSeek inference yields clinical quality scores of 4.58/5.00 at under £7 per 1,000 patient queries—compared to £32 per 1,000 queries for Claude with raw_json at 4.96/5.00.

**Figure 3:**
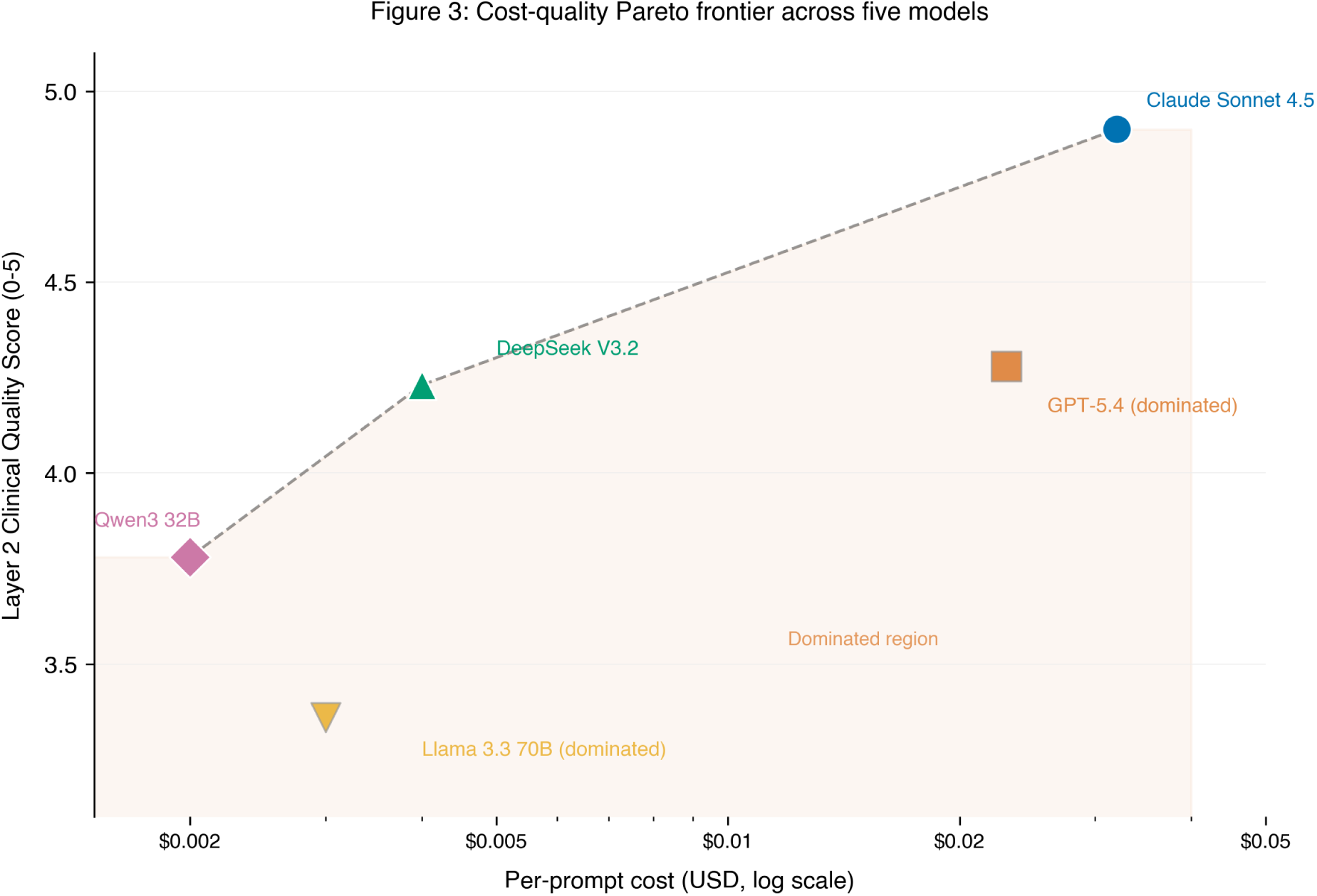
Cost–quality Pareto frontier. Serialisation format provides a second optimisation axis beyond model selection.

## 4 Discussion

### 4.1 Context-Dependent Serialisation: A New Paradigm for Clinical FHIR Processing

The central contribution of this study is the demonstration that FHIR-to-text serialisation format is not a neutral preprocessing step but a significant, controllable determinant of clinical LLM quality. The effect is substantial (0.24 points on a 5-point clinical quality scale, p< 10^−33^), replicable across data conditions, and—critically—context-dependent. There is no single optimal serialisation format; the best choice depends on clinical task, model capability, and data complexity.

This finding overturns two prior claims. Paper 1 [10] suggested that condensed formats consistently outperform raw JSON for FHIR-based clinical tasks. Our initial Paper 2 analysis incorrectly concluded that serialiser choice “barely matters” (Layer 2 difference <0.1)—an error arising from restricting analysis to Claude alone, the model least sensitive to format variation. The complete cross-model analysis reveals a more nuanced and more useful truth: serialisation strategy is a first-order deployment variable whose optimal configuration varies by context.

The mechanistic explanation for task-dependent optimality rests on the distinct information requirements (see Figure 4) of each clinical task type (see Figure 4):

**Figure 4:**
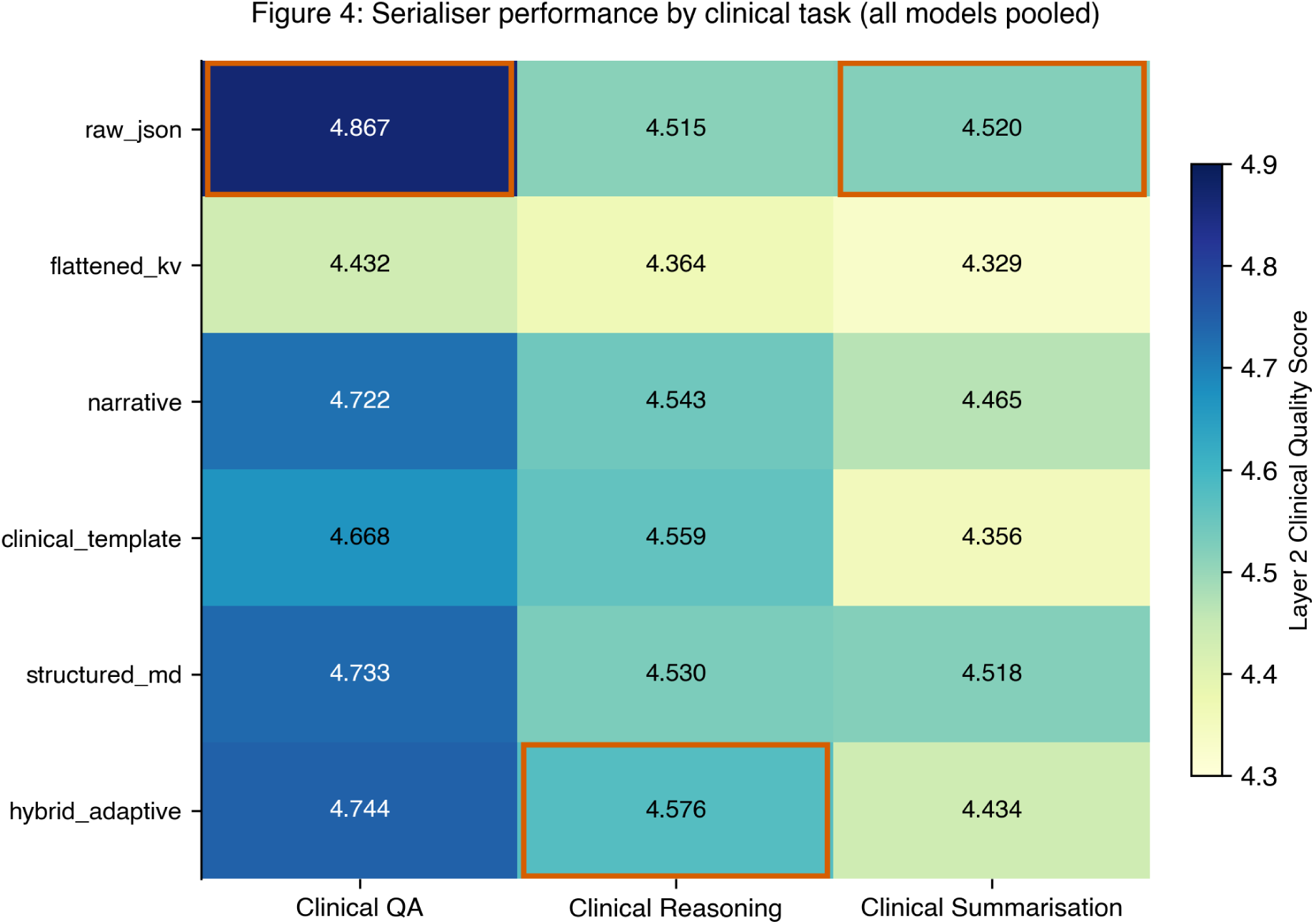
Task × serialiser quality matrix. The optimal serialiser varies by task, motivating task-aware routing rather than static format selection.

**Clinical QA** (factual extraction) requires precise retrieval of specific identifiers: NHS Numbers, SNOMED CT codes, dm+d medication codes, and date-stamped observations. raw_json preserves these tokens in their original form, including system URIs that serve as disambiguation context.

**Clinical reasoning** (drug interactions, risk stratification, care gap identification) requires models to identify relationships between dispersed clinical entities. hybrid_adaptive and clinical_template serialisers pre-organise information by clinical domain, presenting medications alongside their indications and relevant observations in semantically coherent blocks.

**Clinical summarisation** requires hierarchical document synthesis following established clinical communication conventions. structured_markdown provides an inherent hierarchical scaffold via headings and nested lists, which models leverage to produce well-organised summaries.

The convergence with Pator (2026) [1] strengthens this conclusion considerably. That study, conducted independently using different models (open-weight ≤70B), a different task (medication reconciliation), different data (generic FHIR, not UK Core), and a different evaluation methodology (F1 only), arrived at the same core finding: serialisation format significantly impacts clinical LLM performance, and the advantage of specific formats reverses depending on model capability. Two independent research groups, working with non-overlapping model families and clinical tasks, converging on the same phenomenon constitutes substantially stronger evidence than either study alone.

### 4.2 Implications for NHS Deployment: The Adaptive Serialisation Case

The practical significance of context-dependent serialisation is amplified by the finding that model capability inversely correlates with serialiser sensitivity. Claude (highest quality) shows a 0.10-point range across serialisers; Llama (lowest quality) shows 0.39. This interaction produces a deployment paradox: the organisations that most need serialisation optimisation—those deploying budget-constrained models—are precisely those for whom the gains are largest.

An adaptive serialisation layer—a lightweight preprocessing service that routes incoming clinical queries to the appropriate FHIR-to-text conversion based on detected task type—would yield the following gains for a mid-tier deployment (DeepSeek V3.2):

- **Clinical QA workloads**: route through raw_json. Quality: 4.80/5.00.
- **Clinical reasoning workloads**: route through hybrid_adaptive. Quality: 4.69/5.00.
- **Clinical summarisation workloads**: route through structured_markdown. Quality: 4.72/5.00, with 88% token reduction.

The economic case is compelling. For an NHS trust processing 50,000 clinical queries monthly across a mixed workload (40% QA, 30% reasoning, 30% summarisation), adaptive serialisation with DeepSeek V3.2 would cost approximately £200/month whilst maintaining quality above 4.6/5.00. The equivalent quality from Claude with no serialisation optimisation would cost approximately £1,600/month—an 8× premium for marginal quality gains. Recent NHS AI trials demonstrate that even modest cost reductions enable broader adoption across resource-constrained trusts [6, 26].

For NHS trusts operating at scale (*>*100,000 patient interactions per month), the combined effect enables a tiered architecture:

- **Safety-critical tier** (medication reconciliation, A&E decision support): Claude + raw_json. Quality: 4.96/5.00. Cost: ∼£25 per 1,000 queries.
- **Standard clinical tier** (discharge summaries, GP referrals): DeepSeek + structured_markdown. Quality: 4.72/5.00. Cost: ∼£4 per 1,000 queries.
- **Pre-screening tier** (bulk record flagging, population health): Qwen + clinical_template. Quality: 4.18/5.00. Cost: ∼£1.50 per 1,000 queries.

This tiered approach brings LLM-assisted FHIR processing within budget reach of community trusts and GP federations—not only large acute providers. The NHS government mandate [27] and record investment in digital transformation [28] create an enabling environment for such deployments, whilst the NICE evidence standards framework [24] and MHRA AI regulatory strategy [29, 30] provide governance guardrails. The King’s Fund analysis of AI scaling in NHS settings [31] identifies cost and integration complexity as primary barriers—both of which adaptive serialisation directly addresses.

### 4.3 The Ranking Reversal: Cross-National Confirmation

The complete inversion of model rankings between Layer 1 (token-level F1) and Layer 2 (clinical quality) replicates across US Core and UK Core FHIR profiles, two independent synthetic datasets, and two data conditions. The Spearman correlation (*ρ* = −0.90) confirms these are substantively inverted orderings.

The mechanism, which we term the “conciseness trap,” operates through the interaction of response length with metric incentives. Llama’s brevity (mean 724 tokens) maximises F1 precision by minimising tokens absent from ground truth, whilst sacrificing the clinical thoroughness that Layer 2 evaluates.

For the broader clinical NLP community, this carries a methodological imperative: studies reporting only F1, ROUGE, or BLEU for clinical question answering risk conclusions that would invert under clinical expert evaluation. Single-metric benchmarking is insufficient for deployment decisions. Notably, Pator (2026) [1] relied solely on F1 and acknowledged this as a limitation; our multi-layer design demonstrates that serialiser effects observed on F1 do not straightforwardly predict effects on clinical quality.

### 4.4 Robustness and Generalisability

The perturbation experiment provides evidence that findings are not artefacts of idealised synthetic data. Under clinically realistic noise, all model rankings, serialiser rankings, task-specific optimality patterns, and effect sizes remained stable.

However, several limitations constrain generalisability. The evaluation cohort (N=100 patients, synthetic) may not capture the full heterogeneity of NHS production records. The LLM-as-judge methodology carries inherent biases [23]; Claude’s near-ceiling scores (4.90/5.00) may partially reflect alignment between Claude’s generation style and Qwen’s quality preferences as judge. Our sample of five models, whilst representative of major architecture families accessible through Bedrock, excludes models of potential NHS relevance: Gemini, Mistral Large, and domain-specific biomedical models. The benchmark framework is extensible, and complementary evaluations—including work on LLM knowledge of UK public health policy [32] and structured clinical case extraction [33]—demonstrate growing community investment in UK-specific clinical AI evaluation.

### 4.5 Future Directions

Four research priorities emerge: (1) prospective evaluation of the adaptive serialisation routing layer on mixed clinical workloads; (2) extension to additional national profiles (AU Core, EU International Patient Summary); (3) format-aware fine-tuning to reduce sensitivity for mid-tier models; and (4) longitudinal benchmarking across model releases for NHS AI governance, as recommended by the National Commission on AI Regulation [34]. The NHS AI Lab evaluation [35] highlights the need for standardised, reproducible benchmarks to support procurement decisions—a role FHIRBench-UK is positioned to fill. Finally, the flattened_kv format’s consistent underperformance—fragmenting clinical concepts across hierarchical keys—warrants mechanistic investigation into whether this degradation arises from tokenisation artefacts, attention span limitations, or loss of co-reference signals.

## 5 Conclusion

We present FHIRBench-UK, an open benchmark demonstrating that FHIR-to-text serialisation format significantly impacts clinical LLM quality (Kruskal–Wallis H=163.86, df=5, p< 10^−33^; Δ=0.24 on a 5-point scale). This effect is not merely statistically significant but clinically meaningful: in specific model–task–complexity scenarios, switching from a suboptimal to optimal serialiser shifts output quality from clinically unacceptable to clinically adequate. Crucially, the optimal format is context-dependent: raw_json dominates for clinical QA (factual extraction), hybrid_adaptive outperforms for clinical reasoning (drug interactions, risk assessment), and structured_markdown leads for clinical summarisation—whilst achieving 88% token reduction. In 58% of model–task–complexity scenarios, an alternative format outperforms raw_json, with reversals of up to +0.58 points. There is no single ‘best’ serialisation format; the optimal deployment strategy is task-aware routing, not static format selection.

The interaction between model capability and serialiser sensitivity carries direct implications for budget-constrained NHS deployments. Claude Sonnet 4.5 (highest clinical quality) shows only 0.10-point sensitivity to format choice, whilst Llama 3.3 70B shows 0.39—meaning serialisation optimisation is most valuable precisely for organisations deploying lower-cost models where quality margins are tightest. An adaptive serialisation layer—requiring no model retraining, only a preprocessing routing decision—can recover 0.3–0.4 points of clinical quality for mid-tier models at zero additional inference cost. Combined with appropriate model selection, this enables clinical-grade FHIR processing at under £4 per 1,000 queries.

These findings are robust under clinically realistic data perturbation: model rankings, serialiser rankings, task-specific format preferences, and effect sizes all replicate identically across clean and perturbed cohorts. The study additionally confirms the Layer 1–Layer 2 ranking reversal first identified on US Core FHIR data [10], establishing cross-national generalisability across UK Core profiles and reinforcing that multi-layer evaluation—combining automated metrics with clinical quality assessment—is essential for responsible clinical AI deployment decisions. The convergence of our findings with independently conducted work on open-weight models [1] establishes the serialisation effect as a replicable phenomenon across model families, clinical tasks, and evaluation methodologies.

The FHIRBench-UK evaluation pipeline and code are available at https://github.com/JacquelineChong/fhirbench (uk-core branch). The complete dataset—including 992 patient bundles, evaluation cohorts, all prompts, model responses, and judge scores—is archived at Zenodo (DOI: 10.5281/zenodo.21809296).

## Statements and Declarations

## Author Contributions

Jacqueline Chong is the sole author responsible for study conception and design, data generation and curation, software development, experimental execution, statistical analysis, visualisation, and manuscript preparation.

## Data Availability

All data and code are publicly available. The FHIRBench-UK evaluation pipeline and code are available at https://github.com/JacquelineChong/fhirbench (uk-core branch). The complete dataset—including 992 UK Core FHIR patient bundles, stratified evaluation cohorts (clean and perturbed), all 18,000 prompts, model responses, and judge scores—is archived at Zenodo (DOI: 10.5281/zenodo.21809296).

## Ethics Approval

Not applicable. All patient data is fully synthetic. No real patient records were used. The study did not require ethics committee approval.

## Prior Publication

An earlier version of this manuscript was posted as a preprint on medRxiv (DOI: 10.64898/2026.08.05.26359794v1). The present submission contains the same content with formatting adapted for journal requirements.

